# A wearable, steerable, transcranial Low-Intensity Focused Ultrasound system

**DOI:** 10.1101/2023.12.22.23300243

**Authors:** Christopher R. Bawiec, Peter J. Hollender, Sarah B. Ornellas, Jessica N. Schachtner, Jacob F. Dahill-Fuchel, Soren D. Konecky, John J.B. Allen

## Abstract

**Objectives:** Transcranial Low-Intensity Focused Ultrasound (LIFU) offers unique opportunities for precisely neuromodulating small and/or deep targets within the human brain, which may be useful for treating psychiatric and neurological disorders. This paper presents a novel ultrasound system that delivers focused ultrasound through the forehead to anterior brain targets and evaluates its safety and usability in a volunteer study.

**Methods:** The ultrasound system and workflow are described, including neuronavigation, LIFU planning, and ultrasound delivery components. Its capabilities are analyzed through simulations and experiments to establish its safe steering range. A cohort of 20 healthy volunteers received a LIFU protocol aimed at the anterior medial prefrontal cortex (amPFC), using imaging and questionnaires to screen for adverse effects.

**Results:** Simulations and hydrophone readings agreed with less than 5% error, and the safe steering range was found to encompass a 1.8cm x 2.5cm x 2cm volume. There were no adverse effects evident on qualitative assessments, nor any signs of damage in susceptibility-weighted imaging scans. All participants tolerated the treatment well, and the users found the interface effective as the system was capable of accurately targeting the amPFC in all participants. A post hoc analysis showed that “virtual fitting” could aid in steering the beams around subjects’ sinuses.

**Conclusions:** The presented system was successfully used to safely deliver LIFU through the forehead to the amPFC in all volunteers, and was well-tolerated. With the capabilities validated here and positive results of the study, this technology appears well-suited to explore LIFU’s efficacy in clinical neuromodulation contexts.

## 1 Introduction

Transcranial focused ultrasound (tFUS) used at low acoustic intensities is an emerging technique for non-invasive neuromodulation with improved spatial resolution and targeting compared to magnetic or electric non-invasive brain stimulation ^1^. Several studies have been performed using tFUS for non-invasive neurostimulation of targeted brain regions, including of the primary motor cortex and hippocampus ^2^, amygdala ^3^, and thalamus ^4^. The first human transcranial application of focused ultrasound neuromodulation involved stimulation of the frontal cortex applied on 31 patients affected by chronic pain ^5^. Subsequent use of the tFUS technique was described targeting the primary somatosensory cortex of healthy volunteers, in a within-patients, sham-controlled study ^6^. There has been significantly increased interest in the clinical community on the applications of tFUS on neuromodulation, with several recent reviews published in order to summarize the state of the art on this topic ^1,7–10^.

In particular, a systematic meta-analyses on tFUS safety across 33 studies performed in both humans and animals has demonstrated a favorable safety profile ^11^. Early results of measured clinical effects of tFUS in modulating mood and functional connectivity by targeting the right inferior frontal gyrus (rIFG), an area implicated in mood and emotional regulation, have shown promising signal ^12^. Briefly, in a randomized, placebo-controlled, double-blind study, participants who received tFUS reported an overall increase in Global Affect, an aggregate score from the Visual Analog Mood Scales ^13^ indicating a positive shift in mood, which was also objectively indexed via a decrease in functional magnetic resonance imaging (fMRI) resting-state functional connectivity (FC) within resting state networks related to emotion and mood regulation ^12^. These results support that tFUS can be used as a safe and effective means of noninvasive transcranial brain stimulation that has the potential of modulating mood and emotional regulation networks in the prefrontal cortex.

While Transcranial Magnetic Stimulation (TMS) and Transcranial Direct Current Stimulation (tDCS) are FDA-cleared technologies that accomplish similar noninvasive activation of targeted regions within the brain ^14,15^, tFUS may offer a superior alternative to these modalities by providing the ability to target deeper structures than TMS and considerably more spatially selective target regions than tDCS. Just as TMS has shown that repeated high-frequency excitation of the same brain region results in strengthening of synapses through a process known as long-term potentiation ^16,17^, resulting in changes in functional connectivity ^18^, tFUS’ ability to localize energy to smaller and deeper targets opens new opportunities for treating more conditions, improving effectiveness, and reducing side effects.

In this paper we present a wearable system for delivering Transcranial Low-Intensity Focused Ultrasound (LIFU) is presented. The system enables the LIFU energy to be delivered through the forehead via a steerable matrix array transducer to anterior targets in the brain. The steerable beam allows one to deliver focused ultrasound energy to targets within a broad region of prefrontal cortex that are implicated in many psychiatric and neurological disorders. A viable workflow which includes neuronavigation, LIFU delivery planning, and ultrasound delivery components is also presented. Capabilities and performance of the acoustic output of the LIFU probe are experimentally benchmarked to establish the safe and effective steering range. As a validation and proof-of-concept of the design, a cohort of 20 healthy volunteers received a LIFU delivery protocol designed to neuromodulate activity in the anterior medial prefrontal cortex. No adverse effects were evident on qualitative assessments, nor were there any signs of damage in susceptibility-weighted imaging scans, indicating that this system could be safely utilized for future investigative studies.

## 2 Methods

### 2.1 Neuromodulation System

#### 2.1.1 Overview

The prototype system has four main components. They are shown in the block diagram in **Error! Reference source not found.**. The first component is the Openwater Neuromodulation headset, a wearable headset containing a custom ultrasound matrix array. The second component is a neuronavigation system for registering the headset and array to the participant’s MRI data for coarse targeting of the ultrasound beam through array placement. The third component is Openwater Neuromodulation Software, which is used to plan LIFU delivery parameters and determine the signals necessary to precisely steer the ultrasound beam to the participant-specific target location. The fourth and final component is the ultrasound driving electronics, used to apply the electrical signals to the array and deliver the LIFU energy to the target location.

**Figure 1:**
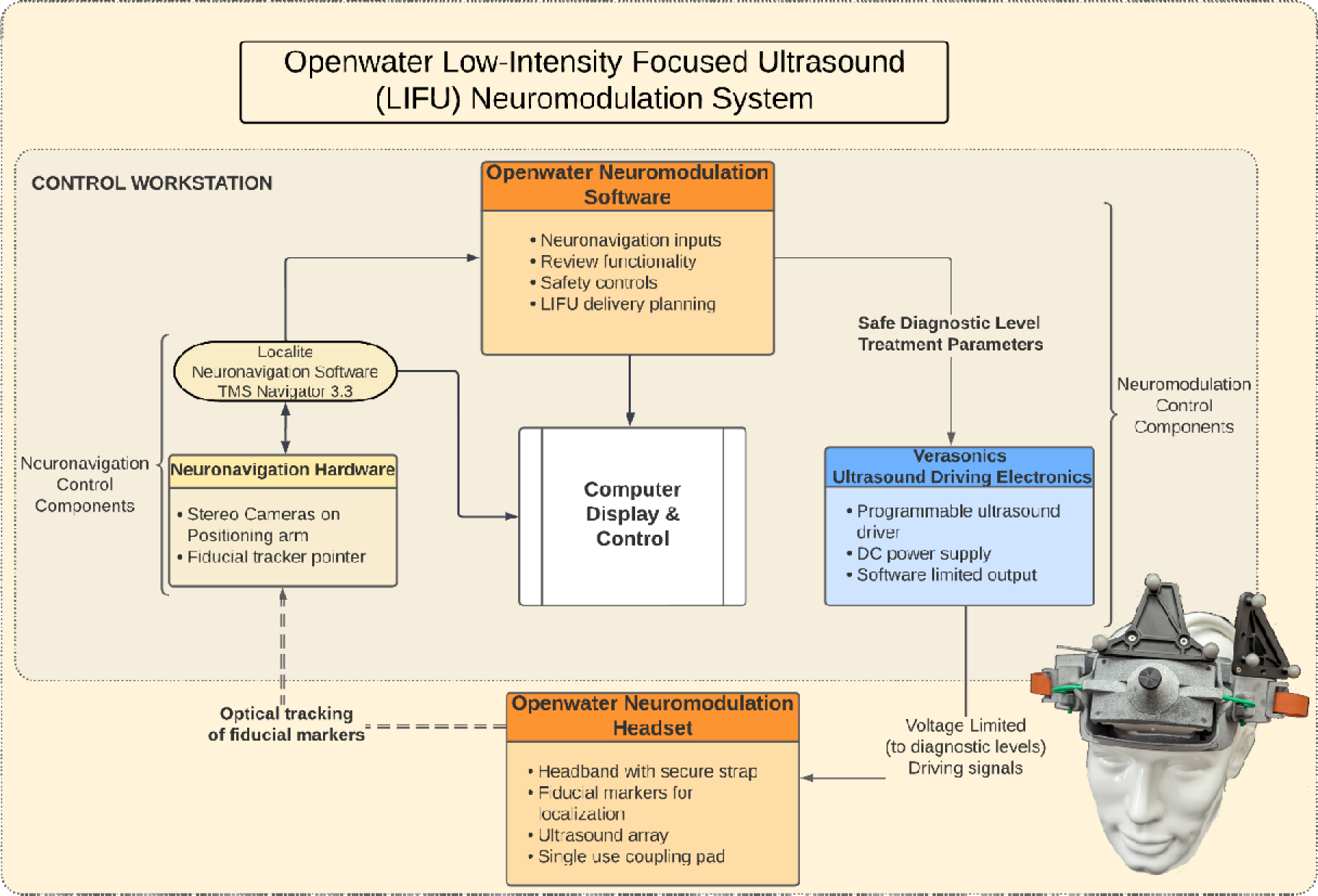
Diagram of the Openwater Low-Intensity Focused Ultrasound Neuromodulation System components.

**Figure 2:**
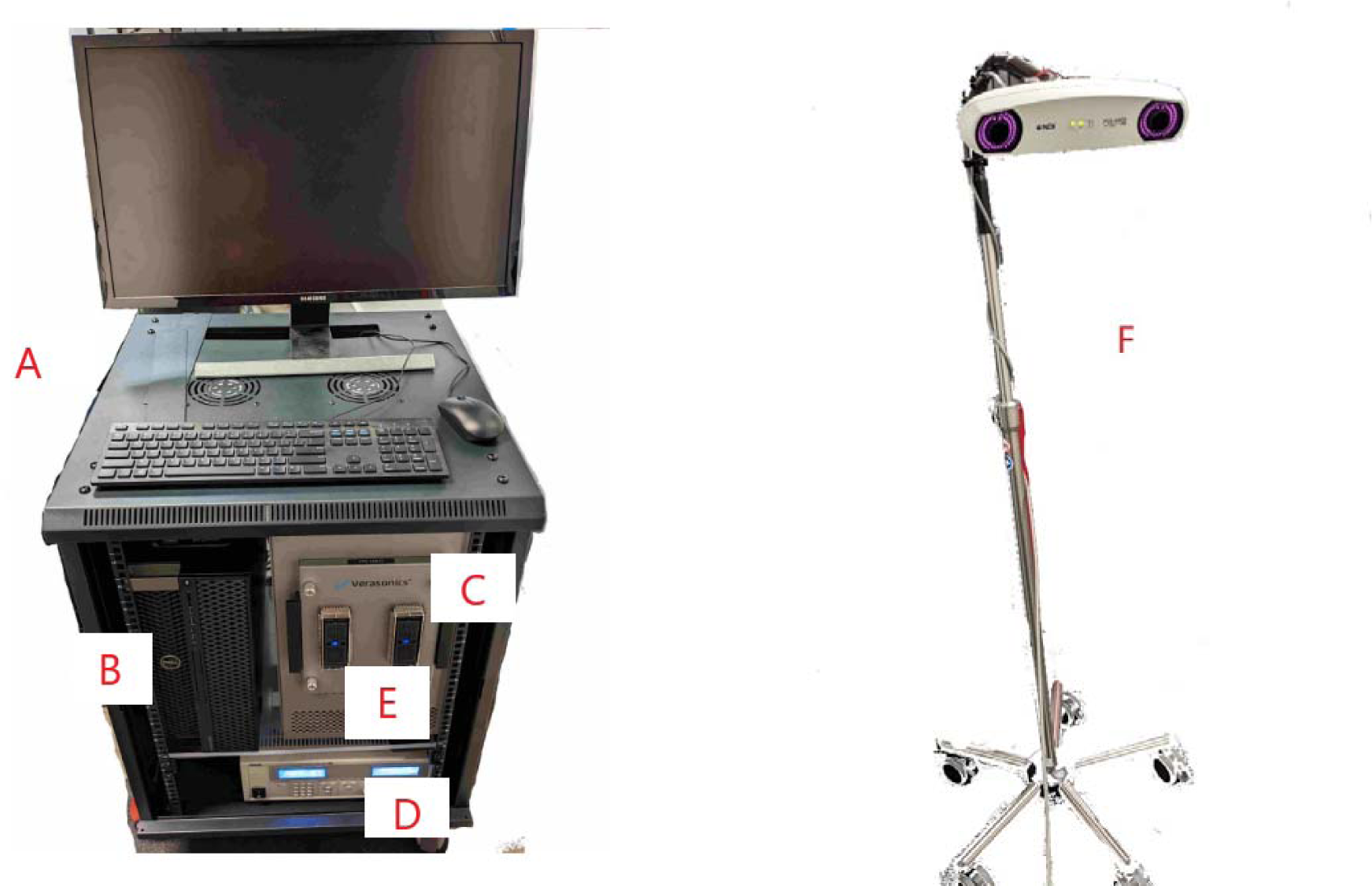
Photo of the system components, (A) Cart with monitor keyboard, and mouse, (B) Computer, (C) Driving electronics, (D) DC power supply, (E) Connector for ultrasound probe, (F) IV pole that holds stereo camera for neuronavigation

#### 2.1.2 Headset

##### 2.1.2.1 Ultrasound transducer array

Openwater’s neuromodulation ultrasound array consists of a 2D array that is designed to be placed and held on the forehead of an individual by a superstructure. The custom 2D array consists of 128 ultrasound elements in 11 rows and 12 columns, with the corner elements not connected. The array is cylindrically focused with a radius of curvature of 50 mm in the horizontal direction. There are two thermistors located in the face of the array ‘s corners to allow temperature monitoring during the tFUS delivery. The elements are square and have a 4.1mm pitch making the surface area approximately 21.5 cm^2^. The array has a 2-meter insulated cable sleeve that carries all of the control and sensing cables from the array to the ultrasound driving electronics. This flexible connection allows freedom of movement for the participant to position themselves comfortably during the session. The cables are routed to a 260-pin Cannon ITT Zero Insertion Force connector through custom PCB boards that have electrical matching components (inductors) soldered to them. These inductors are connected in series to cancel out the capacitive impedance of the transducers array elements at 400kHz.

Three retroreflective spheres with a known geometry are affixed to the array to assist with positioning via the neuronavigation sub-system (discussed in the following section). The 3 spheres have unique distances and serve as triangulation points for the headset and the array, allowing the neuronavigation subsystem to differentiate and track the instruments with high accuracy. The array has a curvature that allows comfortable placement along the anatomical lines of the participant’s forehead.

The ultrasound array’s focus is electronically steerable within a steering range, allowing for flexibility in the placement of the transducer without compromising targeting accuracy. The following two criteria were used to define steering range: First, the intended focus spot must be created without generating grating lobes that have peak pressures greater than 50% of the primary focus; second, the acoustic power that is needed to generate a desired focal spot must not exceed that which would cause the Thermal Index of the Cranium (TIC) to be at or above 3. These limitations mitigate the delivery of ultrasound energy to unintended targets in the brain and allow for an ultrasound duration of 10 minutes, respectively.

**Figure 3:**
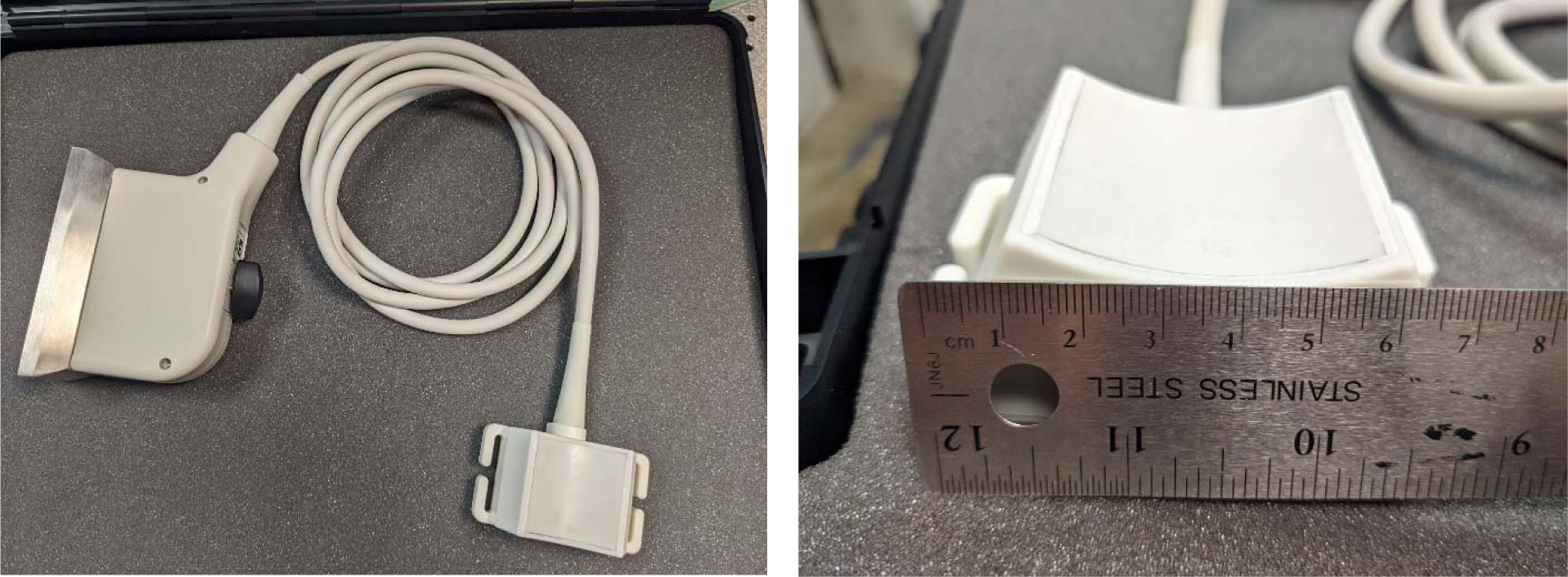
Custom curved-matrix transcranial ultrasound array.

##### 2.1.2.2 Wearable housing and transducer coupling

The wearable component of the system is a 3D printed headset that fits securely on a participant’s head and provides an attachment mechanism for the ultrasound transducer array. The design of the headset allows for the 1-dimensional movement of the probe horizontally along the forehead without repositioning the entire headset. One set of fiducial markers is placed on the headset, which is securely fastened to the participant, for anatomical registration to their MR image. A second set of fiducial markers is placed on the transducer housing and tracks the location of the array when its horizontal position is adjusted.

**Figure 4:**
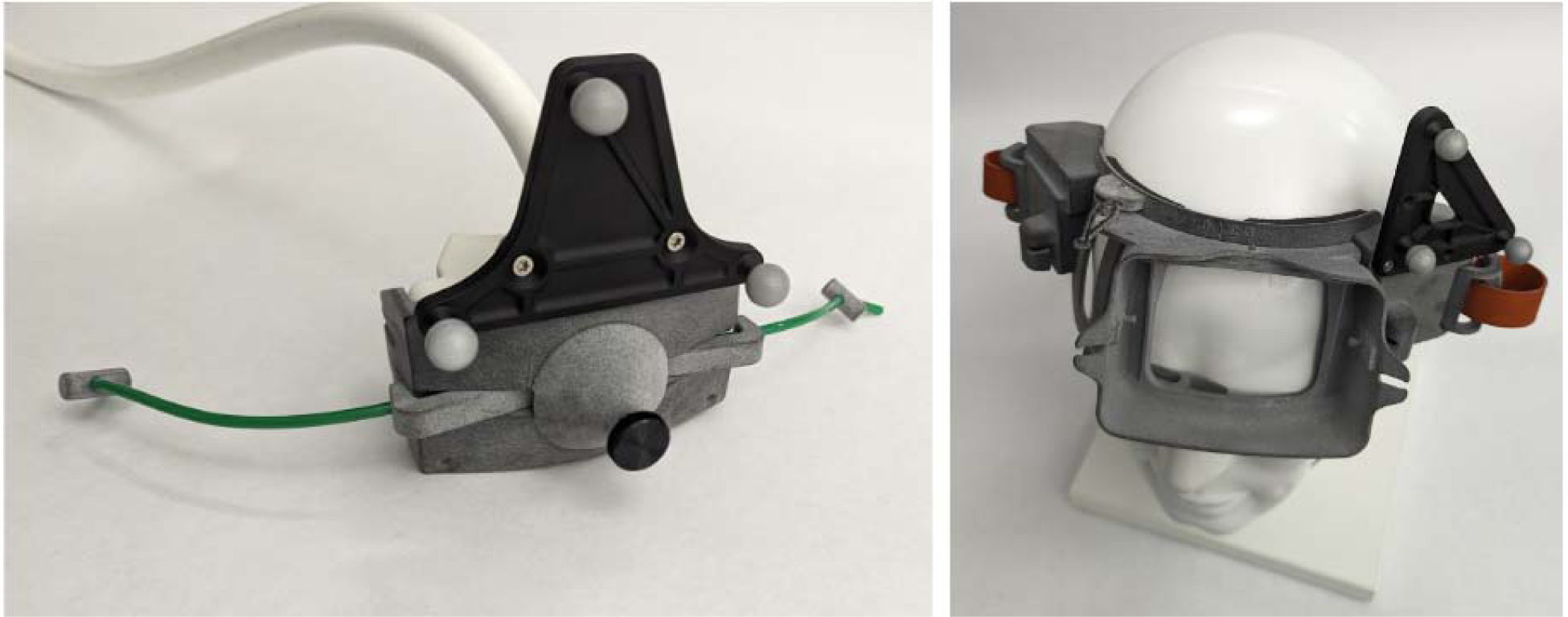
(Left) transducer array in 3D printed housing with fiducial markers. (Right) 3D printed headset with attached fiducial markers on mannequin head, showing opening on the forehead where the transducer array is placed.

The headset has a central positioning support on the lower portion just above the nose that allows it to be placed along the participant’s brow line, between the eyebrows. A custom molded polymer gel pad is placed on the array face and a thin layer of ultrasound gel is placed between the pad and the participant’s skin, assuring that the ultrasound can travel through an acoustically equivalent medium until it reaches the target tissue inside the brain. The polymer gel pad is fabricated using a custom designed plano-convex mold such that when placed on the array the curvature would match a typical forehead. For targeting of the amPFC, the molded polymer gel pad used to acoustically couple the array to the participant was fabricated with a 10° wedge such that it angled the array downwards. The combination of the horizontal movement, the central positioning support, and the wedged coupling pad allowed for positioning of the array within steering bounds of this target.

**Figure 5:**
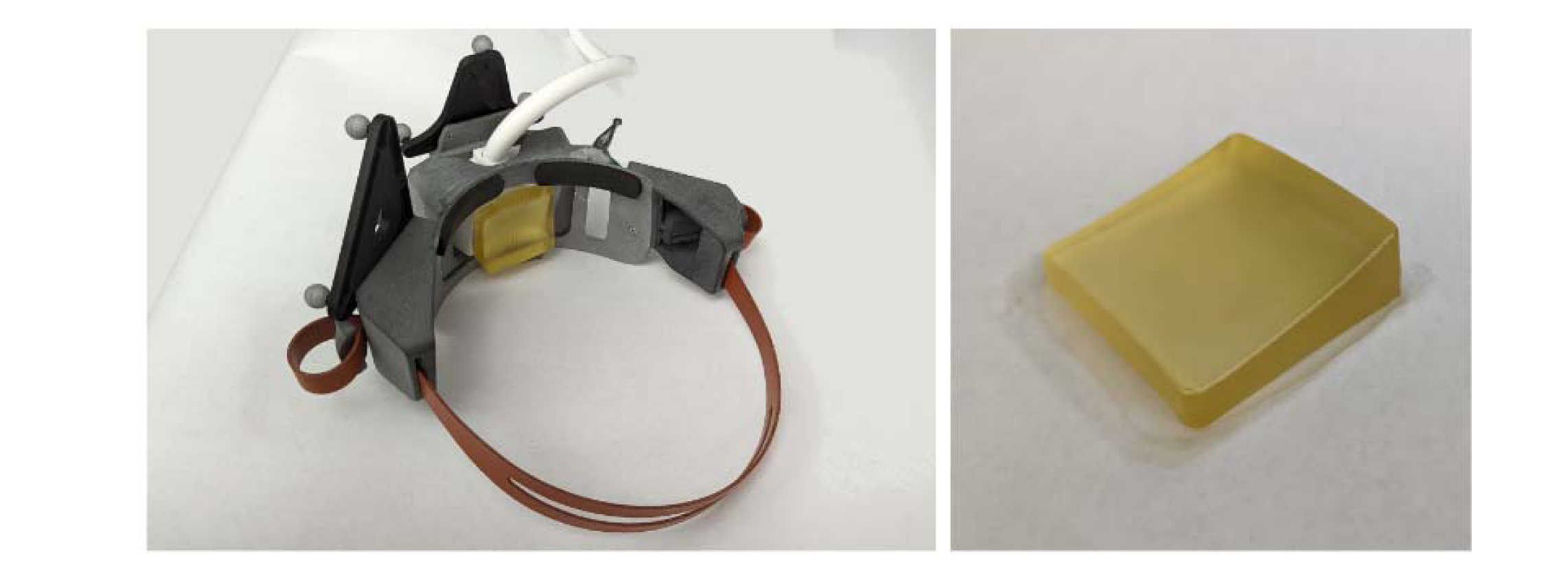
(Left) Headset with transducer in place. (Right) Wedged coupling pad used to couple acoustic energy into the head.

#### 2.1.3 Neuronavigation system

In order to precisely localize the ultrasound transducer array with respect to each individual’s specific anatomy and coregistered medical image, a Localite neuronavigation system (Localite, Bonn, Germany) is used. This neuronavigation system enables the co-registration of the individual’s MR image to their specific anatomy, followed by the registration of the ultrasound transducer array to their anatomy and medical image.

Once the patient’s MRI is coregistered to their anatomical features and mapped to Montreal Neurological Institute (MNI) coordinates, the anatomical target is loaded and appears in the volume. The transducer, attached to its fiducial tracker, is then coregistered into the neuronavigation system software using a custom calibration technique. The transducer is then positioned over the general target location (without a need to be perfectly positioned because the focus is electronically steered), and the Openwater software is used for the rest of the tFUS beam planning and delivery.

#### 2.1.4 tFUS Delivery Planning and Delivery Software

To coordinate the planning and delivery of the tFUS beam, a custom MATLAB software program and GUI was developed. The software interfaces with the neuronavigation and driving electronics subsystems in order to assist with positioning of the headset, compute a beamforming solution for delivering ultrasound from the array to the person-specific target, simulate the delivered acoustic field while ensuring its parameters are within acceptable limits for safety and efficacy, and configure, initiate, and control ultrasound delivery. The ultrasound planning and delivery sequence is outlined in the flow chart of **Error! Reference source not found.**. The details of each step are described below.

**Figure 6:**
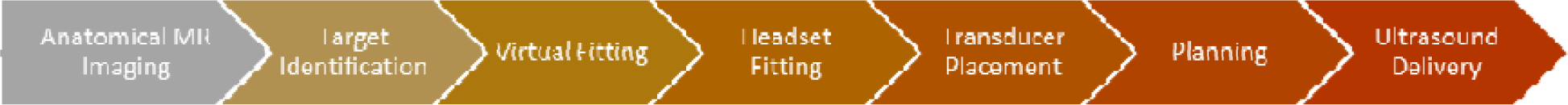
Ultrasound planning and delivery sequence.

**Figure 7:**
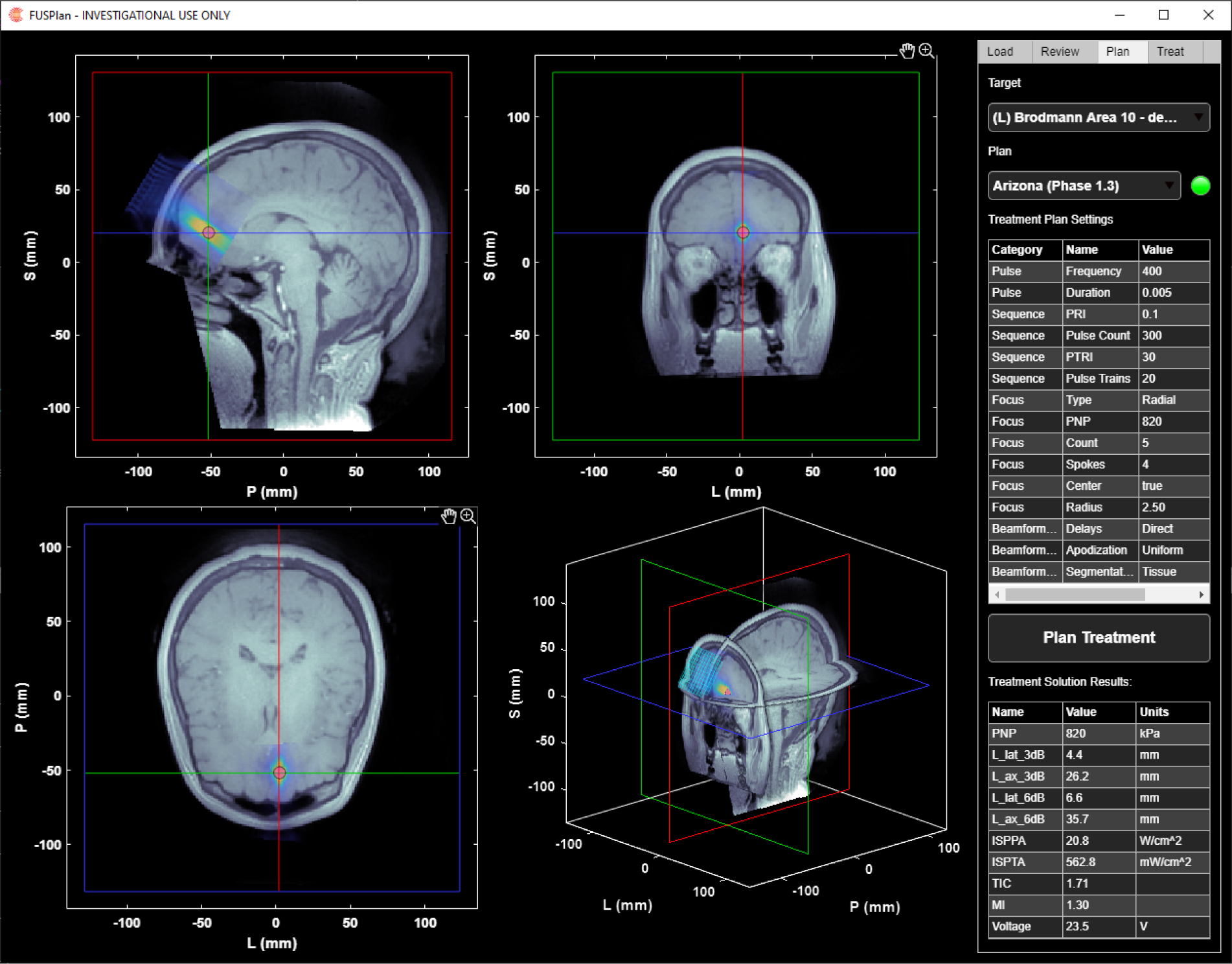
Openwater planning software.

##### 2.1.4.1 Virtual Fitting

In order to aid the positioning of the array, the software recommends an optimal location. The algorithm optimizes the placement by minimizing the distance between the array and the target while ensuring that the target is within the acceptable steering range of the array (defined in section 2.3.3 Establishing Steering Limits), and by minimizing the presence of air-filled sinuses located between the array and the target that will block the ultrasound delivery. The optimization is performed as follows. Once the person-specific target location has been defined in the person’s anatomical MR scan (as described in the prior section), the MR volume and target location are imported into the Openwater software. The outer surface of the skin is automatically segmented via simple thresholding to provide a surface map in polar coordinates. Next, a series of candidate positions for the ultrasound array are defined. For each position, the corresponding subsection of the skin surface is fit to a plane to determine the approximate normal vector to the skin surface. The known transducer geometry is then virtually placed relative to the skin surface, offset to account for the wedged shape of the coupling pad. In each such candidate position, the MRI data between each element of the ultrasound array and the patient specific target are analyzed as a “pyramid” of rays (**Error! Reference source not found.**). This analysis examines two things: the position of the target relative of the transducer to determine how much steering will be needed to direct the ultrasound beam to the target, and the presence and extent of very dark voxels along the rays which are indicative of the sinus cavities (and thus expected to effectively block acoustic waves).

Detection of the sinuses is performed as follows:

1. Trace rays with equal step sizes from each element to the target.
2. Normalize the brightness of the ray so that the average of the first 1mm of all rays is set to 0 (air the head), and the 90^th^ percentile of the last 2mm of all rays is set to 1 (white matter).
3. For each normalized ray, determine if any voxels fall below 10% of the white matter brightness.

**Figure 8:**
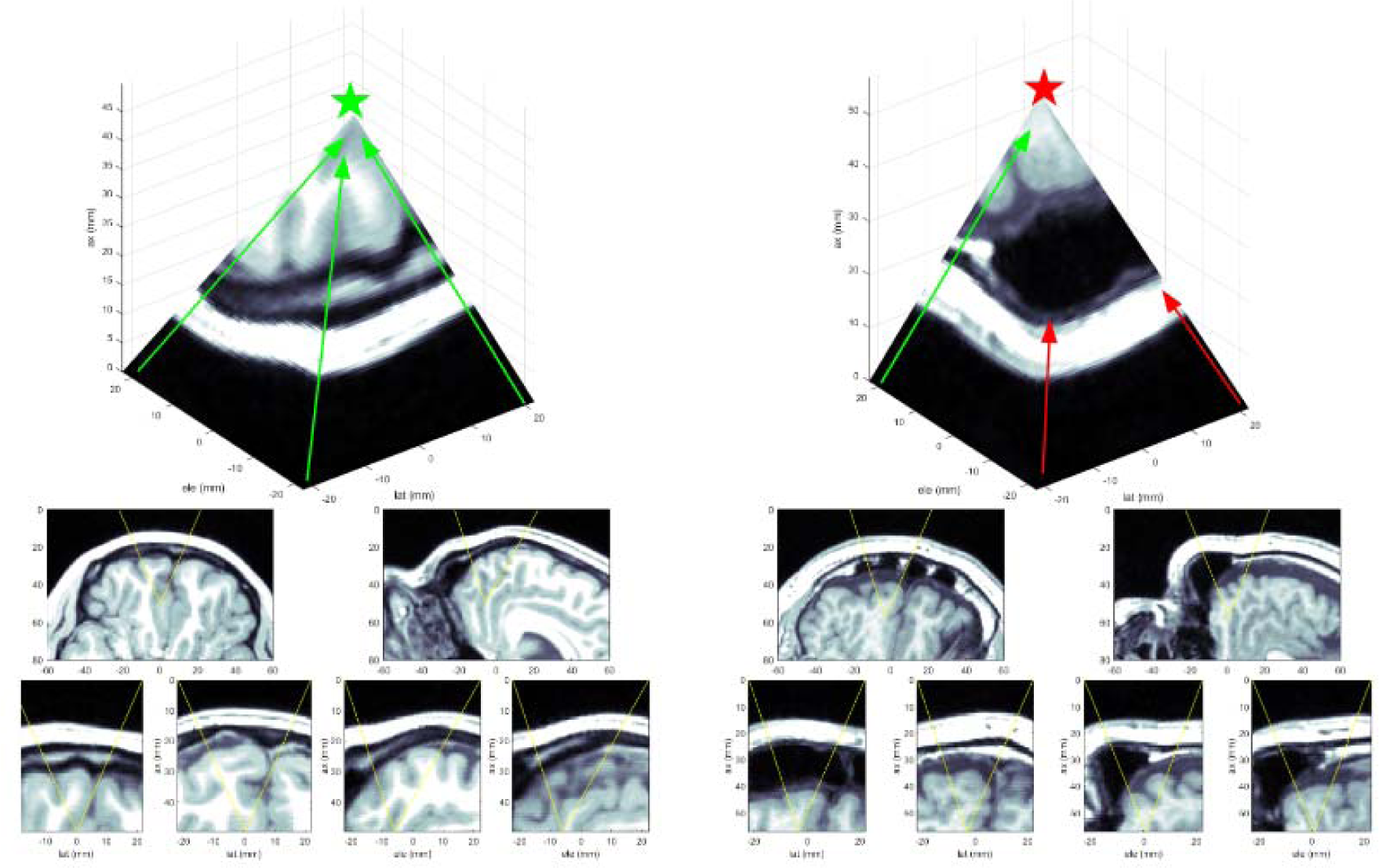
Blocked Sinus comparison. Left: An unobstructed path to the target. Right: a path obstructed by the sinuses.

Once the steering and number of blocked elements has been determined for each position, candidate positions whose steering is outside the valid steering range for the probe are excluded, and then the “optimized” position is selected as the position within the lowest decile of blocked elements that is closest to the target. This position, along with maps of blocked elements and steering validity, are presented to the user, overlaid on the detected skin surface, to provide a visual guide for where to place the probe.

**Figure 9:**
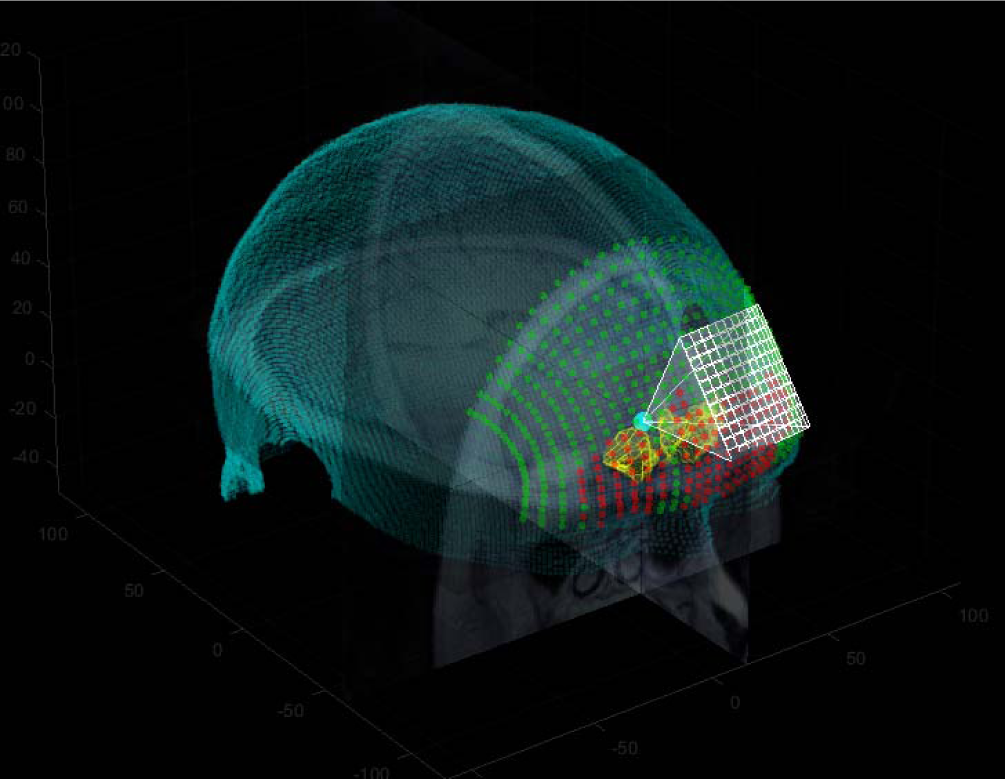
Visualization of Sinuses and probe placement.

##### 2.1.4.2 tFUS beam Planning

Once the participant is fit with the headset, the software tracks the position of the array via the neuronavigation system and provides near-real-time feedback on the steering and number of blocked elements, as were computed in the headset placement planning stage. Once the array is secured in place, the software imports the final array position and loads the requested ultrasound parameters. Next, the MRI volume and target are transformed into the transducer array’s frame of reference, which will be the simulation space. The imaging volume is then segmented and converted to a map of acoustic parameters. Next, delays are computed to steer the beam to the target(s). Once this initial solution has been generated, it is simulated at a nominal voltage and pressure amplitude using k-Wave to determine the nominal pressure and intensity fields^19^. After simulation, the pressure at the target(s) is compared to the pressure requirements of the plan, and the input voltage and simulated fields are scaled so that the target(s) will achieve the requested acoustic exposure. Finally, characteristic parameters of the acoustic field are determined to ensure that sidelobes and safety parameters are within acceptable limits, and the acoustic fields and computed parameters are displayed interactively on the screen.

For the initial safety and usability study, beamforming was performed neglecting the presence of the skull, assuming that all tissue between the standoff and the target was soft tissue. It was decided that this was the most conservative way to ensure that, regardless of the performance of skull-segmentation methods, no participant could receive more acoustic pressure than the target dose.

##### 2.1.4.3 tFUS Delivery

Once the user has reviewed the targeting solution, the software will initiate a connection to the driving electronics, configure the hardware to deliver the ultrasound, and launch a control module for the user to initiate ultrasound exposure. During an ultrasound sequence, a progress bar shows how far into the sequence the participant is, and the temperature of the array is monitored and displayed; if for any reason the sequence needs to be halted, the user can abort through the control module and/or remove the headset.

#### 2.1.5 Driving electronics

The ultrasound driving electronics allow for sending electrical driving signals to each element of the array individually. It is possible to send an identical waveform to each element with a specified time delay (phase difference), and also possible to modify the waveform to each element individually. The waveforms consist of a 3-level, bipolar square wave signal. The ultrasound driving electronics used in the prototype system is a Verasonics Vantage Ultrasound Acquisition platform (Verasonics Inc, Kirkland, WA, USA). The Verasonics Vantage system is a standard frequency system with a HIFU option that includes an external 1200-W DC power supply (QPX600DP, Aim-TTI, Huntingdon, U.K.) with both DC output channels connected in parallel to allow for greater source current.

### 2.2 Ultrasound Capabilities

Because the system drives a matrix array, it can deliver a wide range of ultrasound parameters. We define a Plan as a specification for an ultrasound sequence, including dose targets, beamforming methodology configurations, and constraints on the on steering and/or acoustic safety parameters. For an individual participant, the position of the transducer, their prior imaging data, and the location of the target location within their head are provided as inputs to the plan and used to generate a Solution, which contains the delays, voltages, and timings that the hardware needs to deliver the specified ultrasound dose for that particular participant. For the system presented here, the options that define a Plan are shown in Table 1: Available Configuration Parameters.

**Table 1:**
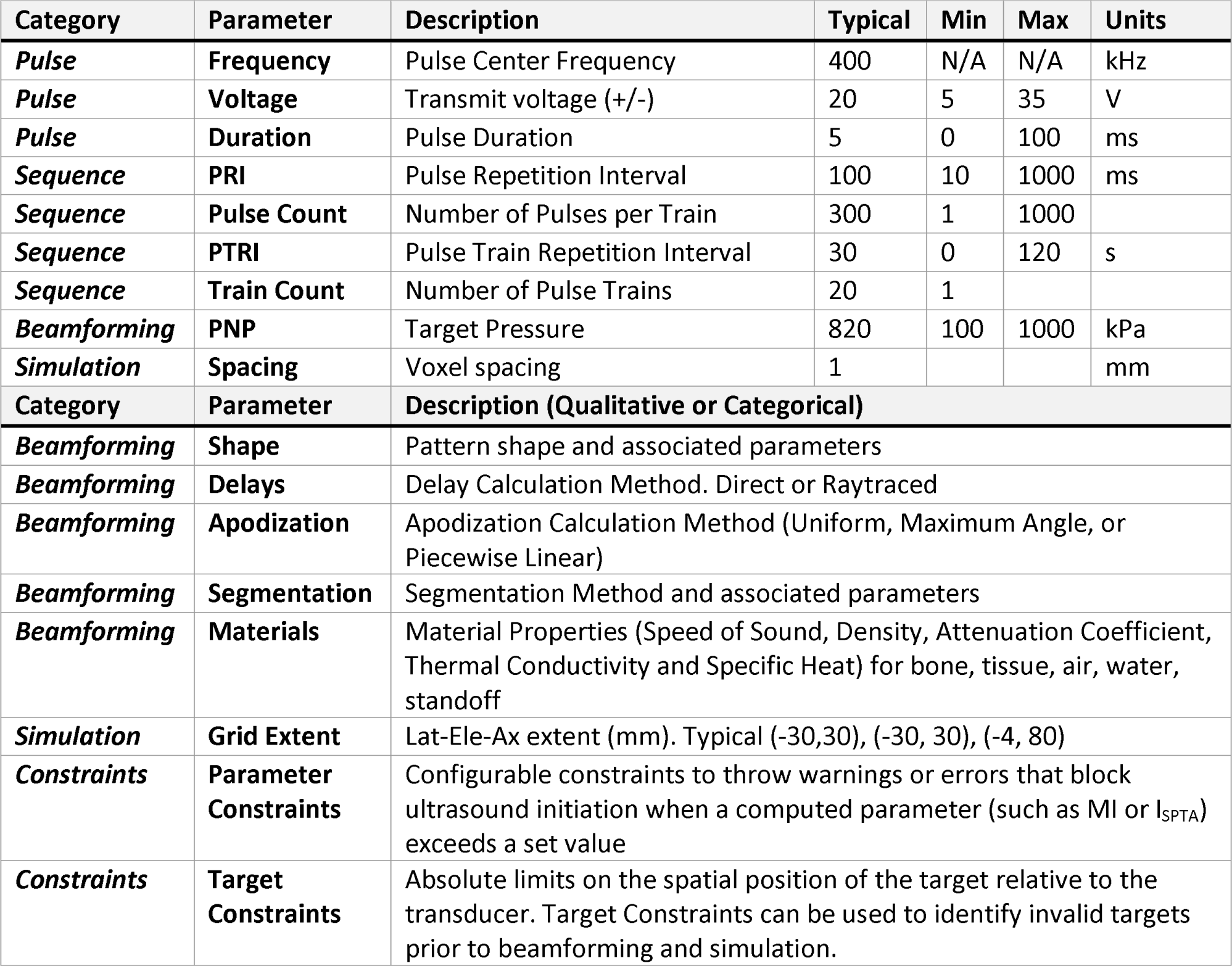
Available Configuration Parameters.

| Category | Parameter | Description | Typical | Min | Max | Units |
| --- | --- | --- | --- | --- | --- | --- |
| **Pulse** | **Frequency** | Pulse Center Frequency | 400 | N/A | N/A | kHz |
| **Pulse** | **Voltage** | Transmit voltage (+/-) | 20 | 5 | 35 | V |
| **Pulse** | **Duration** | Pulse Duration | 5 | 0 | 100 | ms |
| **Sequence** | **PRI** | Pulse Repetition Interval | 100 | 10 | 1000 | ms |
| **Sequence** | **Pulse Count** | Number of Pulses per Train | 300 | 1 | 1000 |  |
| **Sequence** | **PTRI** | Pulse Train Repetition Interval | 30 | 0 | 120 | s |
| **Sequence** | **Train Count** | Number of Pulse Trains | 20 | 1 |  |  |
| **Beamforming** | **PNP** | Target Pressure | 820 | 100 | 1000 | kPa |
| **Simulation** | **Spacing** | Voxel spacing | 1 |  |  | mm |
| Category | Parameter | Description (Qualitative or Categorical) |  |  |  |  |
| **Beamforming** | **Shape** | Pattern shape and associated parameters |  |  |  |  |
| **Beamforming** | **Delays** | Delay Calculation Method. Direct or Raytraced |  |  |  |  |
| **Beamforming** | **Apodization** | Apodization Calculation Method (Uniform, Maximum Angle, or Piecewise Linear) |  |  |  |  |
| **Beamforming** | **Segmentation** | Segmentation Method and associated parameters |  |  |  |  |
| **Beamforming** | **Materials** | Material Properties (Speed of Sound, Density, Attenuation Coefficient, Thermal Conductivity and Specific Heat) for bone, tissue, air, water, standoff |  |  |  |  |
| **Simulation** | **Grid Extent** | Lat-Ele-Ax extent (mm). Typical (-30,30), (-30, 30), (-4, 80) |  |  |  |  |
| **Constraints** | **Parameter Constraints** | Configurable constraints to throw warnings or errors that block ultrasound initiation when a computed parameter (such as MI or $I_{SPTA}$) exceeds a set value | | | | |
| **Constraints** | **Target Constraints** | Absolute limits on the spatial position of the target relative to the transducer. Target Constraints can be used to identify invalid targets prior to beamforming and simulation. |  |  |  |  |

### 2.3 System Characterization

#### 2.3.1 Acoustic Measurements

##### 2.3.1.1 Acoustic field measurements

Characterization of the transducer array’s acoustic focusing abilities were performed to determine the size, shape and intensity of the acoustic focuses generated by electronically steering the array. This enabled determination of the beamforming performance of the array and driving electronics in order to validate electronic steering and simulation accuracy. Measurements were made in a water tank filled with de-ionized, degassed water at room temperature, using a computer-controlled 3D positioning system (AIMS, Onda Corp., Sunnyvale, CA). Calibrated hydrophones (HNR-0500, Onda Corp, Sunnyvale, CA, USA) were used to ensure that measurements were accurate and repeatable.

In order to determine the voltage to pressure relationship for each of the steering locations, pressure field measurements were performed while the array focus was electronically steered. Steered locations included all of the boundaries of the steered volume with intervals not exceeding 5mm from location to location within the volume. These measurements allowed for a 3D interpolation of voltage required for a desired pressure at all of locations within the volume. In order to validate the interpolation, grid spot checking of over 50 locations ensure that the measurements were accurate and repeatable.

##### 2.3.1.2 Acoustic Power Measurements

Acoustic radiation force measurements were performed using a low frequency radiation force balance (Precision Acoustics, Dorchester, United Kingdom). These measurements allowed the determining the acoustic power output of the transducer under various driving conditions. The measurements were taken in de-ionized water at room temperature and the driving conditions were set to match the frequency and apodization of the study parameters but operated in continuous wave mode. Measurements were taken at different driving voltage levels that spanned all of those used during the study. These measurements were then compared to the calculated acoustic power from the simulations.

#### 2.3.2 Simulation Validation

To validate the simulation methods used in the software, simulations of the same signals used for the water tank measurements were performed in both K-Wave and FIELD II ^20^. The simulated and measured pressure fields were visually compared and the −3, −6, and −12dB contours were computed for a sample of steering locations within the steering range of the transducer and overlain on one another to determine how well the spatial pressure fields overlap between methods.

#### 2.3.3 Establishing Steering Limits

Because the shape and amplitude of the focused beam varies with focal location, the transmit voltage is scaled up to compensate for a loss of focal gain when the target is moved away from the nominal focus. For the presented experiments, a target peak-negative-pressure (PNP) at the focus was used to calculate the necessary voltage adjustment. However, using different voltages means generating different surface powers, so thermal heating of the skull (TIC) and other safety parameters become dependent on target location.

A grid of targets, covering −12.5 to 12.5 mm laterally, −20 to 10 mm in elevation, and 40 to 60 mm axially was simulated to cover a reasonably nominal steering range to target the amPFC through the forehead. Within this range, the various beamforming, safety and exposure parameters were calculated for each candidate target location, allowing for the definition of a sub-region of valid targets for which all parameters met acceptability criteria.

#### 2.3.4 Thermal Simulation

Currently, the Openwater system does all of the acoustic simulations with the properties of homogeneous soft tissue. Since heat-related bio-effects are known not to occur until exposures of 1.5°C - 2.5°C above baseline temperature for at least 1 hour, the Openwater system limits heating to 2°C and a maximum possible total ultrasound time of 10 minutes ^21,22^. This is also in line with the latest ITRUSST recommendations which recommend no more than 10 min of ultrasound if the TIC is 3 or above ^23^. To estimate the level of tissue heating associated with ultrasound, k-Wave’s thermal simulation toolbox was used, for both nominal and “worst-case” steering conditions. Because the heating was low, it was validated that the results were effectively the same whether they were obtained by simulating the entire ultrasound duration (with all excitations), or simulating each focal location once individually, and superimposing the differential results for each excitation in the sequence. This second method allowed for rapid investigation of different ultrasound sequences.

#### 2.3.5 Still air and simulated use thermal testing

Thermal testing of the array was performed to ensure that it was able to pass the Still Air and Stimulated use tests outlined in the IEC 60601-2-37 standard. These tests were performed with the same stimulation parameters as the study (5 ms burst, 10 Hz PRF, same waveform apodization), but at a 20% higher driving voltage than used during the study, and no off time during the duration of the tests. The still air test consisted of running the probe at full power while suspended in air with no acoustic coupling of the probe. Measurements were made using thermocouples taped to the face of the probe, thermocouples inside of the probe face, and using a thermal imaging camera (Teledyne FLIR, Wilsonville, Oregon). The thermal study was run for 30 minutes.

For the simulated use test a tissue mimicking material (TMM) phantom was fabricated with a hydrated human skull bone at one side. The TMM was a cube with ∼10 cm sides except for the embedded skull bone. A thin layer (1-2mm) of TMM was poured on the outside of the skull before applying a 1.5mm silicone sheet place on top. The ultrasound array was placed in contact with the silicone through the custom polymer coupling pad with ultrasound gel to closely mimic the use case. Thermocouples were placed in the center of the array above and below the silicone sheet and the ultrasound transducer. The thermal study was run for 20 minutes (double the maximum ultrasound time).

### 2.4 Human participant protocol to assess safety and useability

#### 2.4.1 Study overview

The neuromodulation system safety and useability study used the system to deliver tFUS to human participants and assess system usability, monitor adverse events, and to provide a neuroradiological read of post-delivery susceptibility-weighted images that can probe for vascular micro-hemorrhages. The Institutional Review Board of the University of Arizona approved the experimental protocol. All participants signed an IRB-approved informed consent document before participation.

#### 2.4.2 Participants

Twenty individuals (16F, 4M) participated in this study. Participants recruited for this study were selected to have some degree of repetitive negative thinking, having a score on the Perseverative Thinking Questionnaire (PTQ)^24^ above the 25^th^ percentile. Inclusion criteria also included age 18 – 35, English-speaking, and without any neurological symptoms or symptoms of mania/psychosis. Exclusion criteria included: smoker or use of tobacco products or any form of nicotine daily or nearly daily, history of head injury with loss of consciousness for more than 5 minutes, uncorrected vision and/or hearing impairment, current or history of brain or mental illness judged likely to interfere with testing, including drug and/or alcohol dependence, a diagnosed sleep disorder (e.g., Insomnia), current drug, alcohol, or prescription drug intoxication, history of epilepsy, history of migraines, metal implants in head or face (including permanent dental retainers), history of cardiac problems, current major depressive disorder, a score higher than a 24 on the Beck Depression Inventory II (BDI-II)^25^, and acute suicidal ideation.

Additionally, participants were excluded from the experiment if they were pregnant or unsure if they may be pregnant or have any contraindications for MRI, including severe claustrophobia, non-MRI compatible cardiac pacemakers; implantable defibrillators; aneurysm clips; neural stimulators; artificial heart valves; ear implants; insulin pumps; drug infusion devices; IUDs; magnetic dental appliances; metal fragments or foreign objects in the eyes, skin or body; metal plates, screws, and prosthetics; non-removable metal piercings; tattoos on the head and neck, other certain older tattoos or permanent makeup (eyeliner) using metal-containing inks, some medicated patches, metal implant or other injury or device that is contraindicated for MRI.

Participants were recruited through two methods, the Psychology department introductory psychology participant pool and via flyers in the community. Following consent, participants completed screening questions and the PTQ and BDI-II, followed by MRI scanning, tFUS delivery, and more MRI scanning. The entire study took about two hours to complete.

##### 2.4.2.1 Participant Demographics

Participants’ mean age was 23.6 (SD = 6.21) and 80% were women (16/20, four identified as male), with 80% identifying as white (sixteen White, two Asian, and two Undeclared) and 70% identifying as non-Hispanic (fourteen Non-Hispanic, four Hispanic, two Undeclared). The mean PTQ score was 33.1 (SD = 6.72) and the mean BDI-II score was 11 (SD = 5.58).

#### 2.4.3 Study protocol

##### 2.4.3.1 Baseline Assessments

After the consent process, participants completed baseline surveys assessing their current mood and mindset: the BDI-II, PTQ, Visual Analogue Mood Scale (VAMS)^13^, and the Positive and negative Affect Schedule (PANAS)^26^. These self-report data are not reported here.

Before the ultrasound delivery, participants completed an MRI scanning session, which consisted of a six-minute T1 weighted structural scan, a 3-minute PETRA short TE scan (to assess skull density), and a six-minute resting-state functional scan. Continuous heart rate data via finger photoplethysmography were also collected during the resting-state functional scan. Resting state functional scan and photoplethysmography data are not reported here.

##### 2.4.3.2 Ultrasound Session Parameters

The ultrasound was administered immediately after completion of the baseline MRI. Focused ultrasound targeted the left anterior medial Prefrontal Cortex (amPFC; Talairach coordinates in Table 2). The amPFC is a core hub of the default mode network (DMN) that subserves self-relevant thought ^27^, and was selected for a future study examining tFUS in major depression, a condition characterized by hyperconnectivity within the DMN ^28^. Given the focus on linguistic repetitive negative thought for that future study, the left hemisphere amPFC was favored over the right as the target.

The ultrasound delivery parameters (i.e., frequency, pulse length, and pulse repetition frequency) remained the same throughout the study, but the pressure amplitude was systematically varied in increments to optimize the probability of an effect and ensure safety. In three discrete phases, the maximum pressure and spatial-peak, temporal-average intensity (I_SPTA_) did not exceed diagnostic levels. Additionally, the target location and the dosing scheme were adjusted to target a larger area and increase the likelihood of an effect while maintaining safety and useability.

After the ultrasound device was placed on each participant’s forehead with the custom headset and all parameters were established, participants were instructed to sit quietly and keep their eyes open for the duration of the ultrasound delivery, which was ten minutes. It should be noted that the “virtual fitting” step described in section 2.1.4.1 was developed during the study (in response to the discovery that some subjects had pronounced sinuses), so was not used during headset fitting, but instead used retrospectively to compare the unguided transducer placement to the “optimized” positions.

##### 2.4.3.3 Post-ultrasound Session Procedures

After ultrasound delivery, participants completed another MRI scanning session: up to three six-minute resting state scans and a five-minute Susceptibility-weighted image scan were acquired. Photoplethysmograph continuous heart rate data were also collected during the resting-state functional scans. Participants then completed the same surveys as at baseline in addition to questionnaires assessing their experience with, and safety of, ultrasound delivery: the Toronto Mindfulness scale (TMS)^29^, the Ego Dissolution Inventory (EDI)^30^, and the ultrasound sensation questionnaire. Finally, participants were debriefed of the purpose of the study and knowledge to be gained.

Figure 10 summarizes the human participant study protocol.

**Figure 10:**
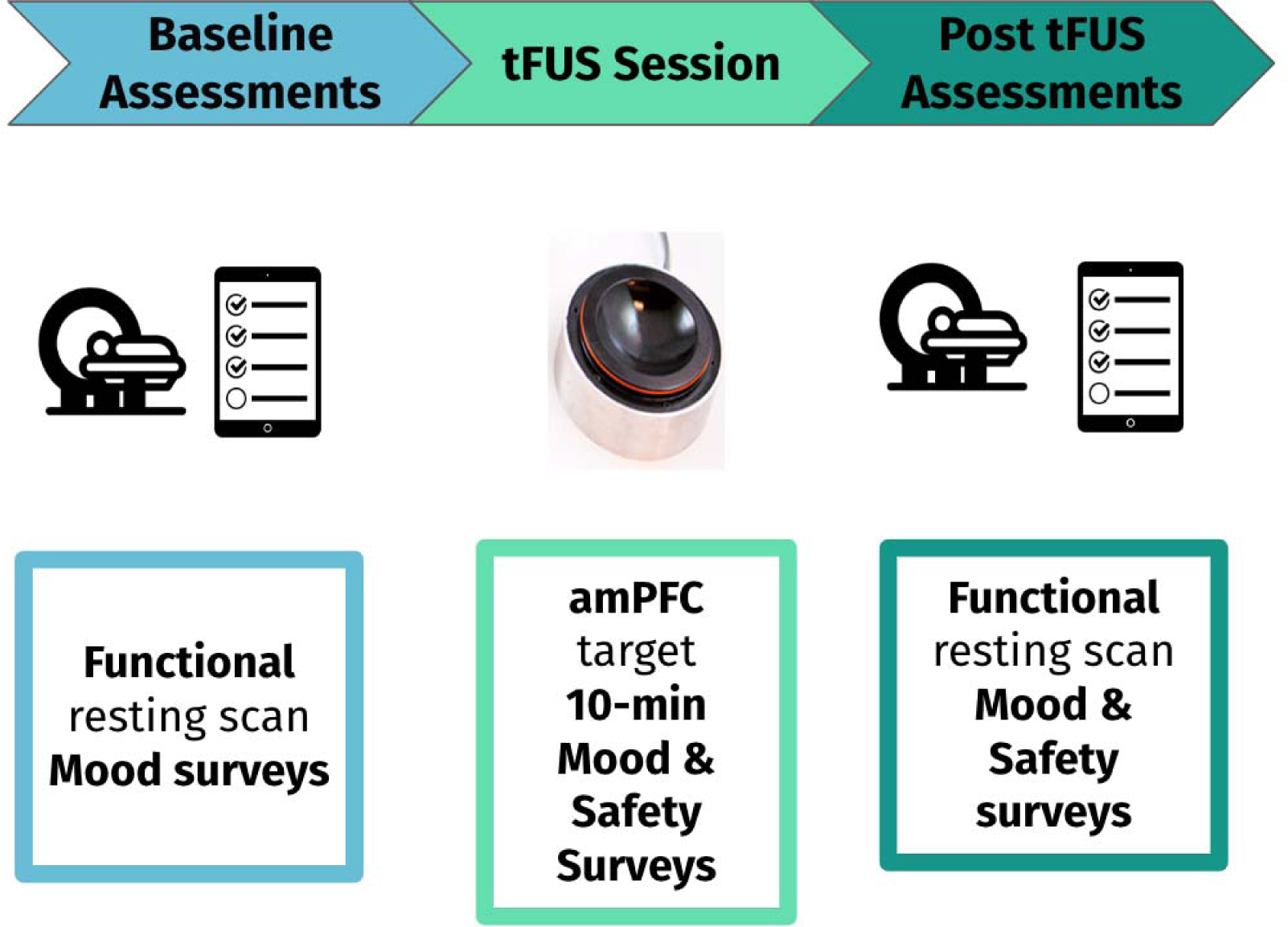
Participants completed one, two-hour session that consisted of three components. First, after consenting, participants completed the following baseline surveys: the BDI-II, PTQ, PANAS, and VAMS. Then, they completed an MRI scan that included a structural TI and functional resting-state scan. After, participants completed a ten-minute ultrasound session targeting the anterior-medial prefrontal cortex. After the ultrasound session, they completed another MRI scan that included a functional resting-state scan and a susceptibility-weighted image to assess safety of ultrasound delivery. Finally, participants completed the same surveys as they did before ultrasound delivery with a few additional measures of their experience after ultrasound and safety: TMS, EDI, and the ultrasound sensation questionnaire.

#### 2.4.4 Magnetic Resonance Imaging Parameters

Two imaging sequences were used to acquire structural MRIs (T1 and PETRA)^31^ that allowed for the identification of the prefrontal cortex target and estimate skull thickness and sinus location for acoustic simulations, and for registration with the neuronavigation system for targeting. All images were acquired on 3 Tesla Siemens Skyra scanner (https://www.siemens-healthineers.com/en-us/magnetic-resonance-imaging/3t-mri-scanner/magnetom-skyra), which is a standard research-grade MRI system.

T1-weighted anatomical images were acquired for registration of the functional scans and to assist in the delivery of the ultrasound. (MP-RAGE; TR = 2100 ms; TE = 2.33 ms; TI = 1100 ms; flip angle = 12; FOV = 256 mm; acquisition voxel size 1 mm × 1 mm × 1 mm).

PETRA anatomical images were acquired for registration of the functional scans and to assist in the delivery of the ultrasound. (TR 1= 5 ms; TE = 0.07 ms; flip angle = 6; FOV = 240 mm; acquisition voxel size 0.9 mm × 0.9 mm × 0.9 mm).

Susceptibility-weighted images (SWI) were acquired to assess safety of ultrasound delivery. (TR = 28 ms; TE = 20 ms; flip angle = 15; FOV = 220 mm; acquisition voxel size 0.6 mm × 0.6 mm × 1.5 mm).

#### 2.4.5 Ultrasound application and parameters

While the simulation results indicated a low likelihood of adverse effects from the ultrasound beams, a conservative stepped approach was taken with volunteers. In Phase 1.1 of the study, consisting of the first 9 participants, a single low peak negative pressure (PNP) amplitude (510kPa) focus was used. Following no adverse events, the PNP for each focus was increased to 650kPa for the 6 participants in Phase 1.2. In addition to changing the focal pressure a new multi-focus approach was implemented that increased the effective area where the ultrasound energy was delivered as detailed below. Following no adverse events in Phase 1.2, Phase 1.3 ultrasound parameters were changed again by increasing the PNP of the pulses to 820kPa, while still keeping the new multi-focus approach of interleaved pulses. This was done to both maximize the ultrasound area and pulse pressure.

The new multi-focus approach (termed interleaved pulses) consisted of targeting multiple foci, one after another (repeating). A radial focus was used (as shown in Figure 13) was used, which positioned 5 foci in a cross-like radial pattern. The distance of the four foci position around the central focus was 5 mm, which corresponded to the width of the focus (−6dB pressure) when steered to the nominal location 50 mm from the center of the face of the transducer array. Each pulse was applied individually but in sequence. This method of pulse interleaving is visually represented in Figure 11. It allowed for spatially distributing the energy delivery in the target region providing a means of increasing the PNP without increasing the I_SPTA_ experienced at any given focus.

**Table 2:** Ultrasound properties used in each phase.

| Phase | N | Target Description | MNI Target | PNP | MI | $I_{SPTA}$ |
| --- | --- | --- | --- | --- | --- | --- |
| **1.1a** | 3 | (L) Brodmann Area 10 | -6, 52, -2 | 510 kPa | 0.8 | 435 mW/cm<sup>2</sup> |
| **1.1b** | 6 | (L) Brodmann Area 10 | - 5, 45, -3 | 510 kPa | 0.8 | 435 mW/cm<sup>2</sup> |
| **1.2** | 6 | (L) Brodmann Area 10 (multi-focus) | - 5, 45, -3 | 650 kPa | 1.03 | <450 mW/cm<sup>2</sup> |
| **1.3** | 5 | (L) Brodmann Area 10 (multi-focus) | - 5, 45, -3 | 820 kPa | 1.3 | <675 mW/cm<sup>2</sup> |

The pulse sequences for each phase are visualized in Figure 11.

**Figure 11:**
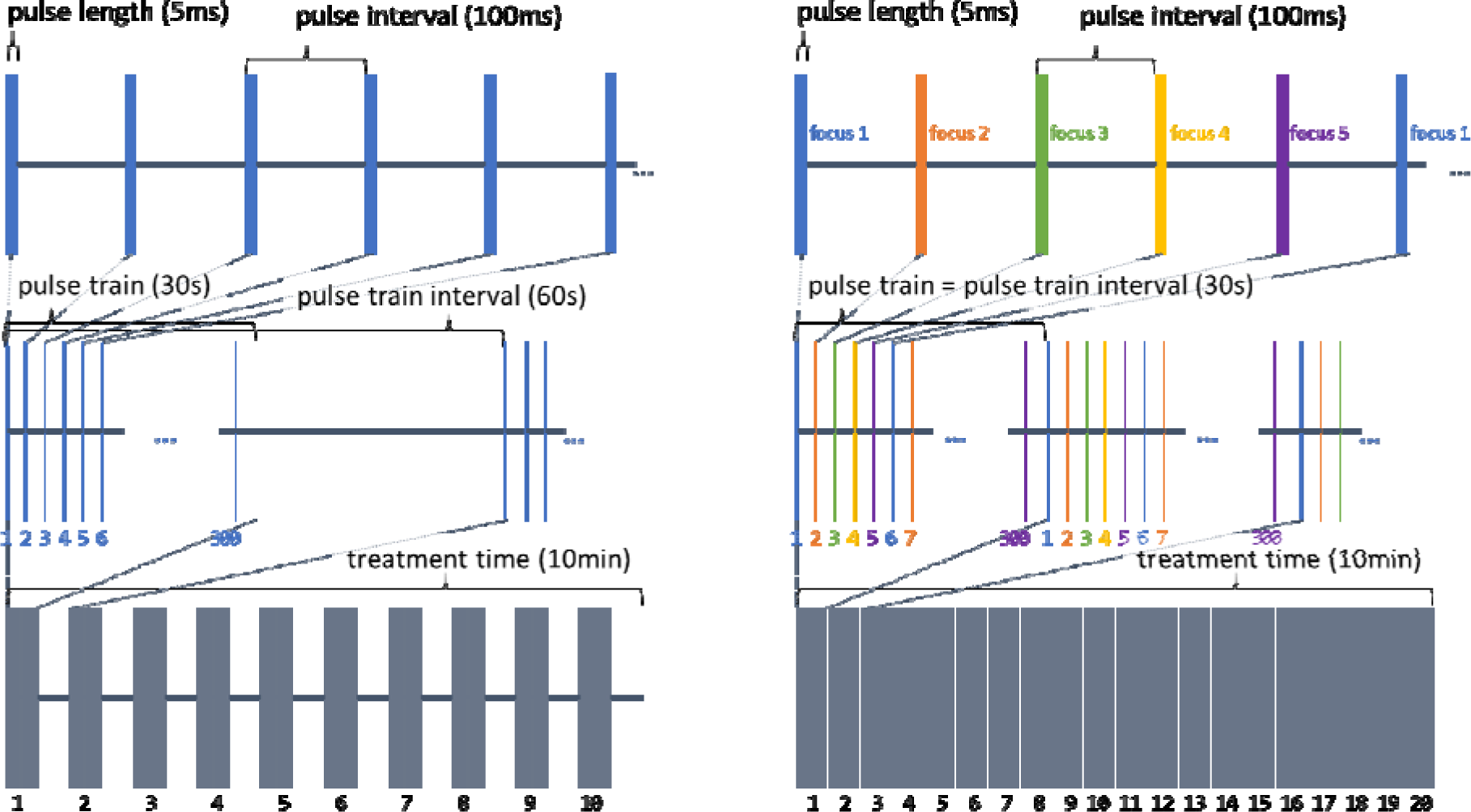
Pulse Sequences. Left: Phase 1.1 Right: Phase 1.2 and Phase 1.3. In Phase 1.1, a single focus is used, with a 30s on-off pattern of pulse trains. In the later phases, the pulses are rastered across 5 partially offset foci, resulting in a longer per-focus PRI but allowing greater spatial coverage and requiring no on-off cycling.

**Figure 12:**
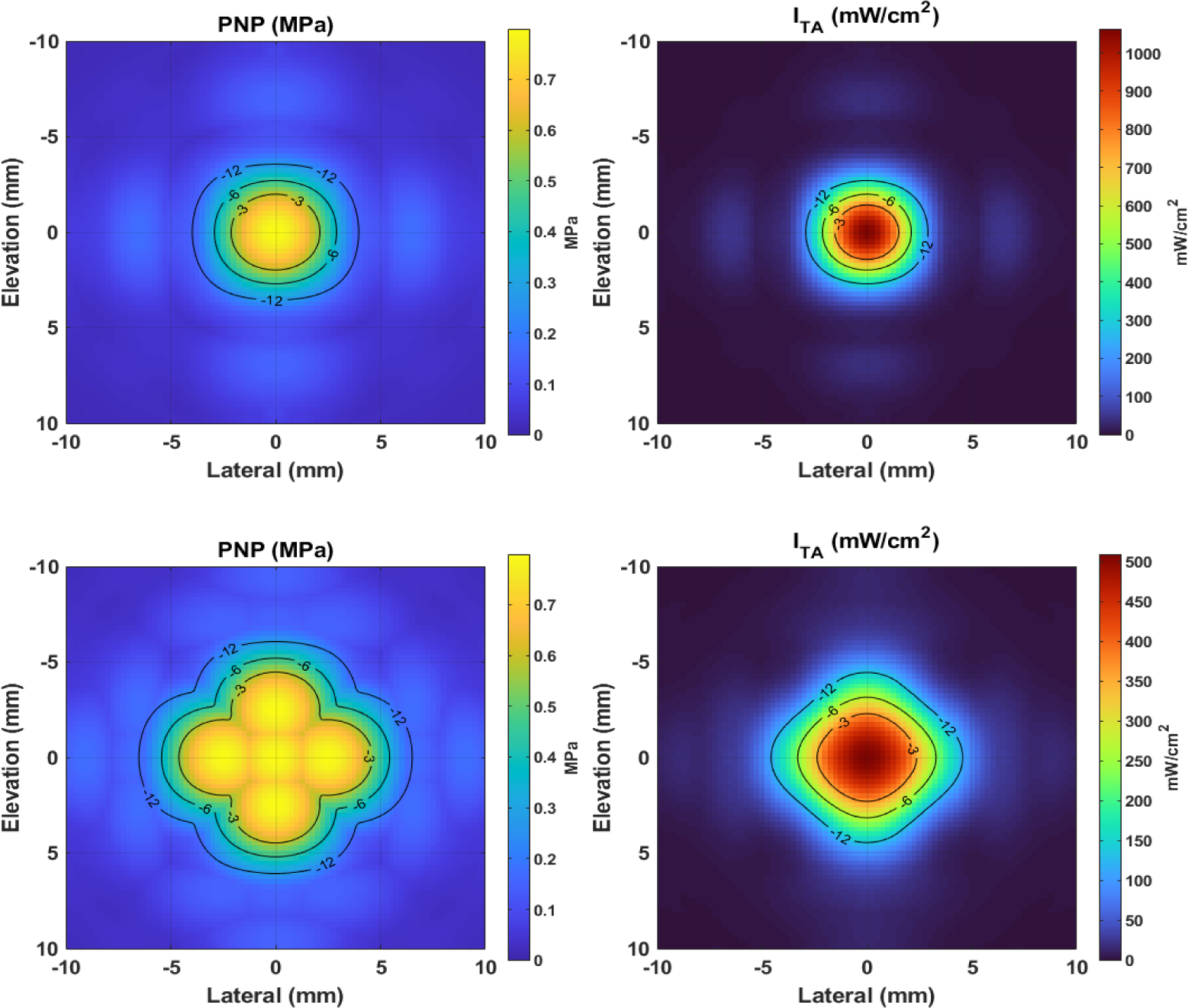
Top: Lateral-Elevation profile of single nominal focus. Bottom: Lateral-Elevation profile of focal pattern used during last phase of safety study. Left: pressure field with −12dB, −6dB, and −3dB contour lines. Right: Temporal average intensity −12dB, −6dB, and −3dB contour lines.

## 3 Results

### 3.1 System Characterization and Validation

#### 3.1.1 Simulation Validation

Cross sections of one of the matched hydrophone scan volumes are shown in Figure 13. The leftmost column shows FIELD II results in the xz and yz planes for a purely water medium, normalized to their global maxima. The second column shows K-Wave results, the third column shows Hydrophone results, and the final column shows the −3, −6, and −12dB contours for each of the three methods. Each slice is also labeled with the Root-Mean-Squared-Error (RMSE) of the pressures between the slice and the hydrophone data as a percentage of the peak. Good qualitative agreement between the contours is seen, and the distribution of pixel values shows little difference in the shape of the beams. Most visual differences are well below the −12dB value, and the RMSE values indicate agreement within 5% between the two simulation methods and the hydrophone measurements.

**Figure 13:**
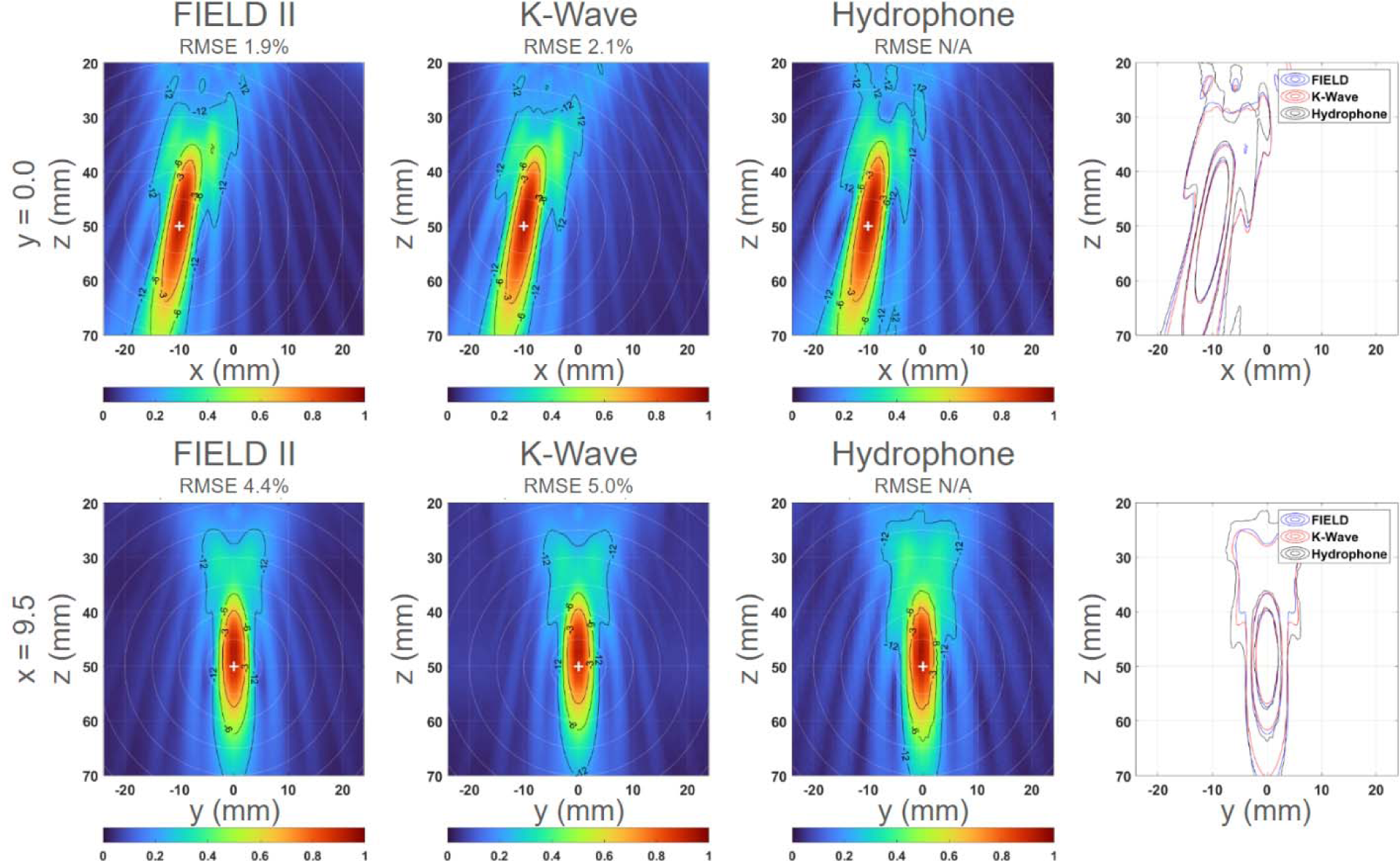
Comparison of simulated and measured beamplots. The top row shows x-z cross-sections, and the bottom row shows y-z cross sections.

#### 3.1.2 Acoustic Output Results

The results of simulating each focal target in the 20 volunteers are shown in **Error! Reference source not found.**, grouped by the three different ultrasound sequences used in each phase.

**Table 3:** Beamforming and safety metrics from k-wave simulation.

| Variable | Phase 1.1 | Phase 1.2 | Phase 1.3 | units |
| --- | --- | --- | --- | --- |
| Number of Volunteers | 9 | 6 | 5 |  |
| Target Steering (Lateral) | $-0.599 \pm 2.3$ | $-2.31 \pm 3.2$ | $0.119 \pm 2$ | mm |
| Target Steering (Elevation) | $-4.11 \pm 7.1$ | $-4.47 \pm 4.1$ | $-1.14 \pm 5.4$ | mm |
| Target Steering (Axial) | $50.7 \pm 3.6$ | $52.3 \pm 2.6$ | $48 \pm 4.2$ | mm |
| Peak Negative Pressure (PNP) | 510 | 650 | 820 | kPa |
| Pulse-Average Intensity ($I_{SPPA}$) | 8.04 | 13.1 | 20.8 | W/cm<sup>2</sup> |
| Time-Average Intensity ($I_{SPTA}$) | 201 | 327 | $508 \pm 46$ | mW/cm<sup>2</sup> |
| Lateral-Elevation (-3dB) Beamwidth | $4.32 \pm 0.26$ | $4.29 \pm 0.19$ | $8.64 \pm 0.54$ | mm |
| Axial (-3dB) Beamwidth | $24.5 \pm 2.6$ | $25.3 \pm 2$ | $23 \pm 2.9$ | mm |
| Lateral-Elevation (-6dB) Beamwidth | $6.43 \pm 0.42$ | $6.28 \pm 0.49$ | $10.7 \pm 0.66$ | mm |
| Axial (-6dB) Beamwidth | $34.3 \pm 1.8$ | $35.3 \pm 1.2$ | $33.1 \pm 3.3$ | mm |
| Thermal Index (TIC) | $0.325 \pm 0.02$ | $0.536 \pm 0.036$ | $1.48 \pm 0.22$ | |
| Mechanical Index (MI) | 0.806 | 1.03 | 1.3 |  |

#### 3.1.3 Steering results

TIC was the limiting factor on the ability of the system to steer to targets with higher pressure, because the limited steerability of the array required scaling up the transmit voltage (and thus emitted power) to compensate for decreased sensitivity. The results of the volunteer targets are shown in **Error! Reference source not found.**, overlain onto the background TIC volume for the most aggressive ultrasound plan (phase 1.3). The results indicated that no targets had exceeded a TIC of 2.5 (no points are located in the yellow regions), which means all participants were well within the TIC<3.0 safety limit for a 10-minute ultrasound. Across all 20 participants, target locations were distributed as follows: lateral (x): −0.9 ± 2.6 mm, elevation (y): −3.5 ± 5.8 mm, axial (z): 50.5 ± 3.7 mm.

**Figure 14:**
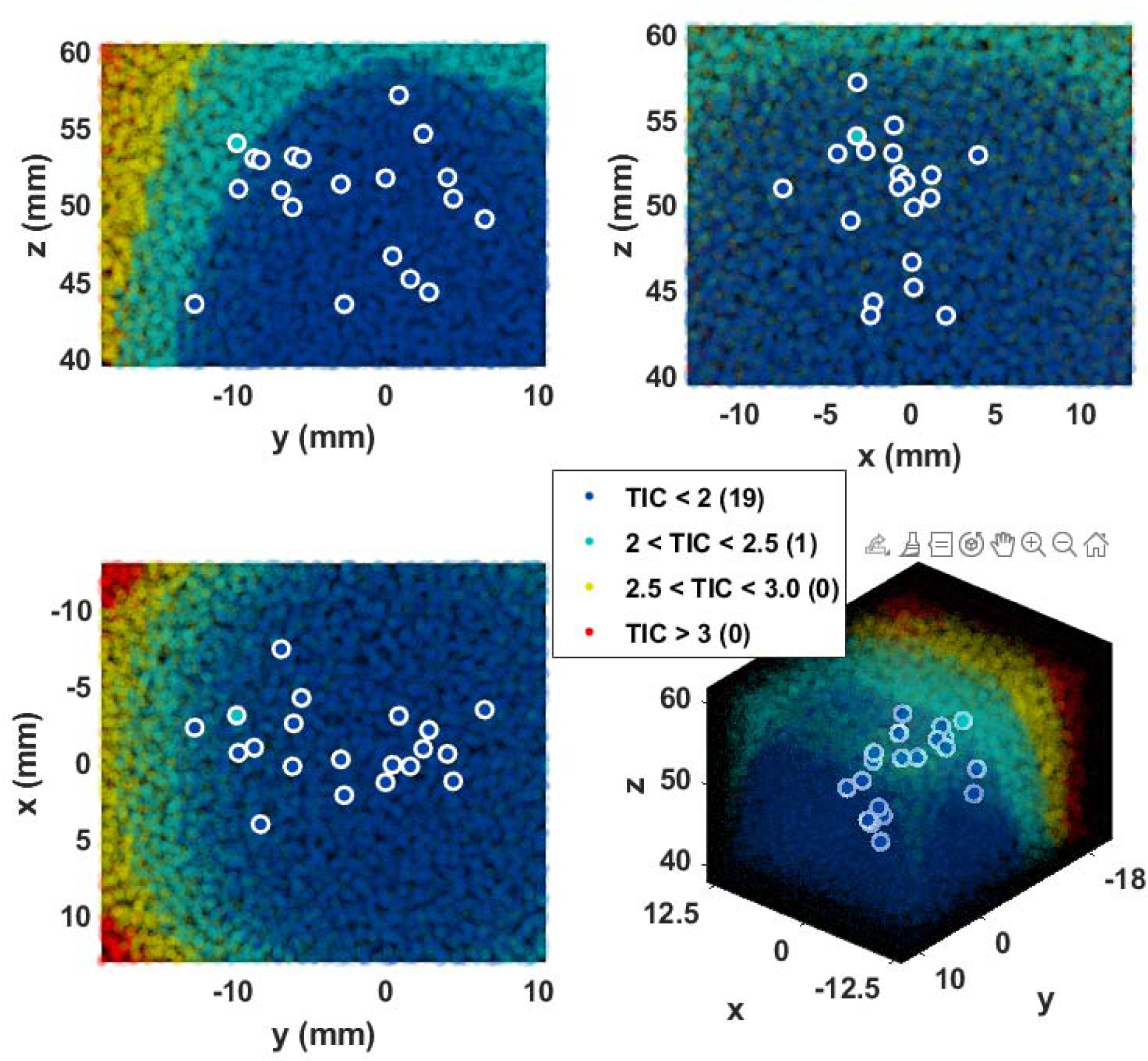
Steering volume represented by TIC value.

#### 3.1.4 Still Air and Simulated Use Thermal results

Thermal testing of the array was performed to ensure that it was able to pass the Still Air and Stimulated use tests outlined in the IEC 60601-2-37 standard. These tests were performed with the same stimulation parameters as the study but with a higher driving voltage and no off time during the duration of the tests. These tests both passed indicating that in the worst case (highest driving voltage) scenario, there was no risk of thermal damage to any participant with the probe in contact with their skin.

### 3.2 Preliminary Safety/Feasibility Study

#### 3.2.1 Usability and tolerability

Operators of the device reported that the system was easy to use and that the interface was effective at guiding the operation of the device. It was also reported anecdotally that all the participants found the headset tolerable and some of the participants even found the sessions themselves relaxing.

#### 3.2.2 Absence of Adverse Events

No serious or subjective Adverse Events (AEs) were reported from subjects. In addition to monitoring AEs, a “sensation” questionnaire was administered to assess potential sensations subjects may experience (on a scale from 0 – 10, with 0 being no sensation to 10 being a high amount of sensation) from the ultrasound, including: itching, heat/burning, tingling, vibrating/pulsing, sound, tension, and pain.

For all sensations, the modal response was 0 (no sensation), as shown in Table 4. Importantly, for aversive sensations very high rankings were absent: Burning/Heat max=5, Pain max=2). For pain specifically, the three individual reports of pain (ratings of 1 and 2) were reported to be related to the comfort of the headset. Overall, both subjective and objective safety measures indicate that ultrasound was safely delivered and well-tolerated by participants.

**Table 4:**
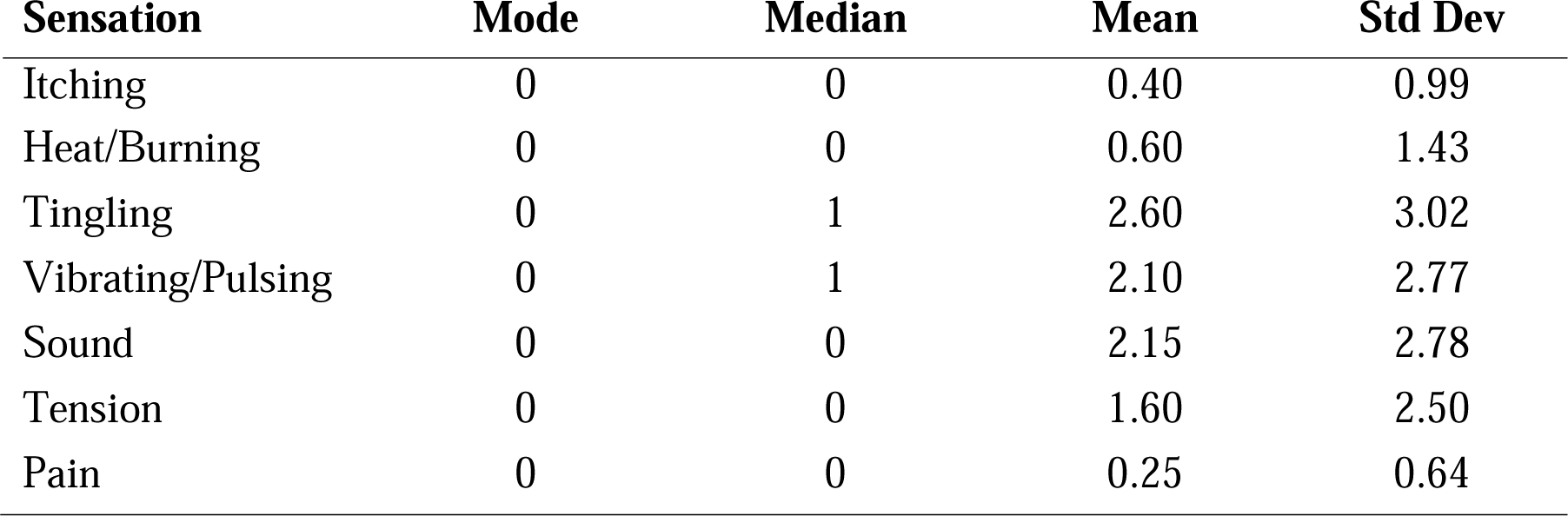
Sensations during ultrasound session.

#### 3.2.3 SWI Imaging

SWI images that were acquired approximately 30 minutes following the completion of the delivery of tFUS were read by two board-certified neuroradiologists. SWI images are sensitive to vascular micro-hemorrhages. All 20 scans were determined to be normal with no findings on SWI, indicating that there were no microhemorrhages.

#### 3.2.4 Sinus Analysis

Although the sinus detection was not used to guide the placement of the probe in the volunteer study, the acoustic paths for the “optimized” transducer position and the “placed” transducer position were compared in **Error! Reference source not found.**. One participant did not have a PETRA scan; thus, the table presents data for 19 of the 20 participants. The position optimization indicated that 8 participants had no sinus blockage of the ultrasound beam. Another 8 subject had some blockage, but the probe could have been moved to locations with no sinus blockage. Out of the three participants where the sinuses could not be completely avoided, two could have had less than 20% blockage of the beam path, and only one had sinuses so large that >30% of the beam would be blocked for any placement of the probe.

**Table 5:**
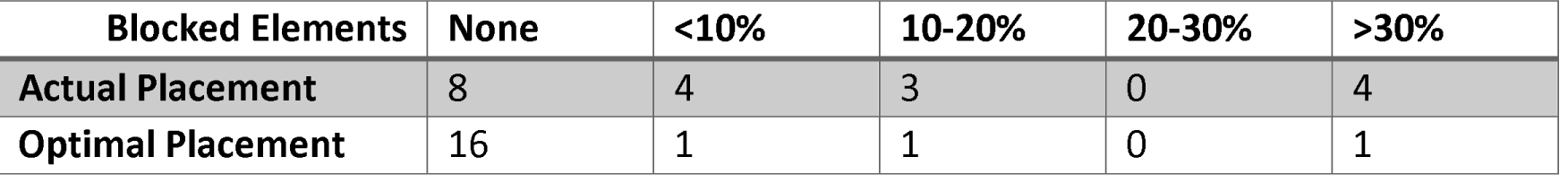
Number of participants with different % of blocked elements for the used and “optimized” transducer placements.

## 4 Discussion

### 4.1 System Capabilities

The system presented here offers unique advantages for precisely guiding LIFU to regions in the front of the brain. It combines a means of rapid positioning, using neuronavigation and a mechanical slider on the headset, with precise control of the targeting, through beamforming, allowing for quick probe placement without compromising accuracy. The probe’s design allowed for a relatively coarsely-diced and modestly-sized matrix array, whose steerable volume is approximately 3×3×3 cm, and it can reach targets up to 6cm deep in most locations beneath the forehead. While a fixed-focus probe is simpler to operate and model, it has drawbacks. To reach the target, it needs precise positioning with a variable entry angle and distance from the head. This would require a bulky, external arm mechanism, making it unwearable. This two-pronged approach (physical and electronic steering) allows the array to be positioned in a way that avoids obstacles like sinuses, making it more versatile and maneuverable. And, although not used in this preliminary study, the system retains access to more advanced beamforming techniques, including element-wise adjustment of apodizations and delays allowing for aberration correction. The small size of the array also means that the headset can be comfortably worn for the duration of the ultrasound without bulky fixturing.

### 4.2 Volunteer Study

The study found successful placement of the headset and delivery of transcranial ultrasound in all participants. Out of 19 participants analyzed, it was shown that only one had sinuses so large that it would not have been possible to reposition the probe to avoid blocking >30% of the elements. For all participants, there were no reported adverse events or findings on the post-delivery SWI images. Because the volunteers in this study were chosen to not have the type of abnormal neural connectivity associated with certain types of depression, it remains to be tested whether targeting the amPFC with this system will reduce depressive symptoms in those afflicted. Future work will also explore whether aberrant default-mode network activity can be suppressed with tFUS in participants likely to have it, as well as assess the magnitude of such participants’ clinical response to tFUS neuromodulation therapy.

### 4.3 Conclusions and Future Work

This neuromodulation system safety and useability study had LIFU delivered transcranially to the amPFC of twenty participants. This study assessed system usability, monitored adverse events, and provided a neuroradiological readout of post-delivery susceptibility-weighted images that can probe for vascular micro-hemorrhages. There were no adverse events or imaging findings, all participants tolerated the sessions well, and the users thought the interface was effective.

With the capabilities presented here and positive results of the feasibility/safety testing, this technology appears well-suited to examine transcranial focused ultrasound’s effect in the clinic. The next clinical work will explore the feasibility of the system to neuromodulate targets in participants with clinical depression.

## Data Availability

All data produced in the present study are available upon reasonable request to the authors

## Acknowledgements

This study was funded, in part, by an investigator-initiated grant to JJBA by Openwater. The authors wish to thank Scott W. Squire and Heidi Elledge for their invaluable assistance with obtaining MRI scans. The authors are also indebted to Jessica Andrews-Hannah and Achal S. Achrol for discussions on target selection and refinement. CRB, PJH, and SDK are full-time employees of Openwater.

